# Effect of Tai Ji and/or Qigong on Patients with Stable Chronic Obstructive Pulmonary Disease A protocol for meta-analysis and systematic review

**DOI:** 10.1101/2023.09.21.23295879

**Authors:** Hongliang Liu, Ningchang Cheng

## Abstract

**Background:** Chronic obstructive pulmonary disease (COPD) is a global health problem with high morbidity and mortality. Tai Ji and Qigong are traditional Chinese mediative movements, benefit COPD patient’s physical and mental health.

**Methods:** We searched the following twelve databases Web of Science, EBSCO, Medline, EMBASE, Scopus, PubMed, PsycArticles, Psychology and Behavioral Sciences Collection, PsycInfo, CINAHL, Cochrane Library online and Clinical trials from inception to July 2023. Any RCTs managed with Tai Ji and/or Qigong on stable COPD were eligible without age, publishment language and comparison management restrict. Outcome measures comprised pulmonary function, the incidence of acute exacerbation, 6WMD, chronic pain, physical and/or cognitive function, and any assessment of people QoL.

**Results:** Our research will update evidence summaries and provide a quantitative and standardized assessment of the effect of Tai Ji and/or Qigong on patients with stable COPD.

**Conclusion:** Our research will generate the latest evidence for determining whether Tai Ji and/or Qigong is equivalent to conventional PR.

## 1. Introduction

Chronic obstructive pulmonary disease (COPD) is one sort of chronic respiratory disease characterized by progressive and partially reversible airflow limitation and presented with chronic cough and dyspnea. Acute exacerbation of COPD (AECOPD) is defined as periodic deterioration of respiratory symptoms, resulting in the need for hospitalization or urgent care, worsen patient’s lung function and quality of life even to death. COPD is a global health problem with high morbidity and mortality which led to almost 3.3 million deaths yearly and imposes a substantial economic burden that would cost worldwide economic trillions of dollars in future thirty years [1]. The prevalence of COPD was 8.6%, and 13.7% among individuals over forty years old in China [2], and increased gradually caused by population aged and long-time exposure under environment with high risk (tobacco, smoke or PM2.5).

Pharmacological treatment was long-termly considered as cornerstone of COPD treatment, nowadays more insights were focus on early treatment, disease stabilization and prevention of AECOPD. Patient with stable COPD was normally managed by inhaled medications presented as bronchodilators and inhaled corticosteroids (ICSs) [3], however the treatment outcomes were not always satisfactory due to patient’s poor tolerance with pharmacy side effect. Pulmonary rehabilitation (PR) is described as a comprehensive intervention based on a thorough patient assessment followed by patient-tailored therapies that include exercise training, education, and behavior change, designed to improve the physical and psychological condition of people with chronic respiratory disease and to promote the long-term adherence to health-enhancing behaviors [4, 5], and is an essential component of COPD management, would benefit the disease stabilization, and reduce exacerbation. Daily practice of PR included breathing trainings such as pursed lip breathing, yoga breathing, or breathing with computer-aided feedback, and exercise training which aimed to strengthen back, arms, and legs, as well as the muscles used to breathe.

Tai Ji and Qigong (Yijinjing, Liuzijue, Baduanjin et al.) are traditional Chinese mediative movements, originated from thousand years ago, combine with sustained mindfulness, deep diaphragmatic breathing, and gentle movements, applied as the complementary medicine in treating chronic disease, could improve patient’s cardiac and lung function, release chronic fatigue or pain, finally benefit patient’s physical and mental health. Several systematic reviews (SRs) previously revealed that Tai Ji and Qigong are traditional PR, and could improve FEV1, 6WMD, and QoL for patients with COPD [6, 7]. The positive conclusion was seemed certain, however most RCTs included in research were published in Chinese and complained as low quality due to high risk of either randomization or allocation concealment bias. Tai Ji and Qigong are becoming worldwide popularity, more studies with high quality are reported, therefore we tried to figure out a new protocol for study on Effect of Tai Ji and/or Qigong on patients with stable COPD.

## 2. Materials and Methods

### 2.1 Study Protocol

Our research was implied followed by the PROSPERO platform and Cochrane Handbook for Systematic Reviews and Meta-Analysis, and we have registered in PROSPERO (CRD42023428833).

### 2.2 Search Trails

We searched original study in the following twelve databases (details of websites were available in Supplement): Web of Science, EBSCO, Medline, EMBASE, Scopus, PubMed, PsycArticles, Psychology and Behavioral Sciences Collection, PsycInfo, CINAHL, Cochrane Library online and Clinical trials from inception to July 2023. We searched PubMed initially used MeSH terms as “Tai Ji” AND “chronic obstructive pulmonary disease”, or “Qigong” AND “chronic obstructive pulmonary disease”, and then this search strategy was adopted to each database and run with familiar MeSH terms or key words.

### 2.3 Inclusion Criteria

#### Population

Individuals diagnosed with stable COPD by Global Obstructive Lung Disease (GOLD) or other authoritative diagnostic criteria without age, publishment language and comparison management restrict.

#### Interventions

Any RCTs managed with Tai Ji and/or Qigong were eligible.

#### Comparisons

Conventional therapy and/or PR combined with Tai Ji and/or Qigong or not, or no treatment.

#### Outcomes

Outcome measures comprised pulmonary function (e.g., FEV1, FVC, FEV1/FVC%), the incidence of acute exacerbation, 6WMD, chronic pain, physical and/or cognitive function, and any assessment of people QoL.

### 2.4 Exclusion Criteria

Participants suffering AECOPD, or case report, or conference abstract only, or study with raw data unavailable.

### 2.5 Risk-of-Bias Assessments

The methodological quality for the included study was assessed independently by two researchers (Liu, Cheng) based on Cochrane risk-of-bias criteria [8], and each quality item was graded as low risk, high risk, or unclear risk based on the following criteria: (1) trials were considered low quality if either randomization or allocation concealment was assessed as a high risk of bias, regardless of the risk of other items; (2) trials were considered high quality when both randomization and allocation concealment were assessed as a low risk of bias, and all other items were assessed as low or unclear risk of bias in a trial; (3) trials were considered moderate quality if they did not meet criteria for high or low risk. The items used to evaluate bias in each trial included the randomization sequence generation, allocation concealment, blinding of participants and personnel, blinding of outcome assessment, incomplete outcome data, selective reporting, and other bias. Meanwhile we defined other bias as the different diagnostic criteria on stable COPD in each RCT.

### 2.6 Data Extraction

Two lead researchers (Liu, Cheng) independently screened each study after duplicate remove and extracted the following information: lead author, publication year, country or region of origin, participant characteristics, stage of stable COPD, performance details, frequency and duration of intervention, and outcome targeted. Disagreements were resolved by consensus.

### 2.7 Statistical Analysis

The association of Tai Ji and/or Qigong with stable COPD was assessed, and Tai Ji and/or Qigong was separately compared with conventional therapy or no treatment group. We performed meta-analysis to calculate risk ratios (RRs) or absolute risk differences (ARDs), and 95% CIs by the Mantel-Haenszel statistical method using Revman version 5.4 (Cochrane Collaboration). A random-effects model was used to pool the data, and statistical heterogeneity between summary data was evaluated using the *I*^2^ statistic. Sensitivity analysis was performed when *I*^2^ > 50%, aimed to exclude low-quality studies, or trials with characteristics different from the others. We specified subgroups based on the exercise frequency of Tai Ji and/or Qigong (≥ 4 or < 4 times a week), and the place which program was taken [9] (home-based, community-based, or hospital-based). All tests were 2-tailed, and *P* < .05 was considered statistically significant.

## 3. Discussion

The benefits to COPD patients from PR were certainly considered, included the improvement of dyspnea, health status, and exercise tolerance across all grades of COPD severity [5]. Traditional PR (Tai Ji, and Qigong) remained the first option of PR in China, with lower economic burden and more effectiveness than conventional PR. Liu W reported Tai Ji combined total body recumbent stepper (TBRS) exercise respectively decreased St George’s Respiratory Questionnaire (SGRQ) total score and improved QoL in mild to severe stable COPD people [10]. Polkey MI reported twelve weeks exercise cessation in bronchodilator-naive COPD patients brought a clinically significant difference in SGRQ emerged favoring Tai Ji (vs. PR based on standard UK practice) [11]. A RCT revealed that the changes in CAT scores before and after the intervention were significantly different between the two groups (Qigong vs. aerobic exercise using a cycle ergometer) [12]. However, whether traditional PR is equivalent to conventional PR is in debate, traditional PR is becoming worldwide popularity owing to strengthen participant’s awareness of availability and benefit, and low-cost during exercise.

Tai Ji and Qi gong are mind controlled physical exercises, benefit both physical and psychological health status. A SR that included twenty-three studies tentatively supported that Tai chi effectively reduced anxiety and depression, and improved general mental health compared to non-mindful exercise [13]. A SR identified seventeen randomized trails implied Tai Ji and Qigong promoted cognitive function in the elderly directly/indirectly through enhancing physical function [14]. Back to COPD people, a small RCT reported that there were between-group improvements favoring Tai Ji (vs. ordinary treatment) in Centre for Epidemiologic Studies Depression (CESD), and Patient-Reported Outcome Measurement Information System (PROMIS) – fatigue, then supported Tai Ji as a potential modest role in improving cognitive-emotional health and function [15]. A pilot study indicated Yijinjing training contributed to improving the ability of emotion regulation, despondency/distress management and QoL [16].

SRs and Meta-analysis previously indicated positive result of Tai Ji and/or Qi gong on physical and psychological health of COPD patient, however the results were blamed on low quality of included trails. Here we try to find a new protocol for SR and meta-analysis, first, we set a proper target on search trails, second, we defined other bias as the different diagnostic criteria on stable COPD in each RCT, third, we specified subgroups based on the exercise frequency of Tai Ji and/or Qigong, and the place which program was taken.

### Supplement

- Web of Science (https://www.webofscience.com/)
- EBSCO (https://www.ebsco.com)
- Medline (https://www.medline.com/)
- EMBASE (https://www.embase.com/)
- Scopus (https://www.scopus.com/)
- PubMed (https://pubmed.ncbi.nlm.nih.gov/)
- PsycArticles (https://www.apa.org/pubs/databases/psycarticles)
- Psychology and Behavioral Sciences Collection (https://www.ebsco.com/products/research-databases/psychology-behavioral-sciences-collection),
- PsycInfo (https://www.apa.org/pubs/databases/psycinfo),
- CINAHL (https://www.ebsco.com/products/research-databases/cinahl-database)
- Cochrane Library online (https://www.cochranelibrary.com/)
- Clinical trials (https://clinicaltrials.gov/)

## Data Availability

If the manuscript reports pilot data held in a public repository, include URLs, accession numbers or DOIs. If this information will only be available after acceptance, indicate this by ticking the box below. For example: All files are available from the pubmed database.

## Abbreviations

RCTs=: randomized controlled trials
6WMD=: 6-minutes walking distance
QoL=: quality of life
CIs=: confidence intervals
CAT=: COPD assessment test

